# Exploratory Study to Characterise the Individual Types of Health Literacy and Beliefs and Their Associations with Infection Prevention Behaviours amid the COVID-19 Pandemic in Japan: A Longitudinal Study

**DOI:** 10.1101/2023.04.02.23287895

**Authors:** Mao Yagihashi, Michio Murakami, Mai Kato, Asayo Yamamura, Asako Miura, Kei Hirai

**Affiliations:** Division of Scientific Information and Public Policy, Center for Infectious Disease Education and Research, Osaka University, Suita, Osaka, Japan; Osaka University Graduate School of Human Sciences, Suita, Osaka, Japan

## Abstract

**Background:** During a global infectious disease pandemic such as the coronavirus disease 2019 (COVID-19), individuals’ infection prevention/risk-taking behaviours are likely to differ depending on their health literacy and beliefs regarding the disease. To effectively promote infection prevention behaviours, it is necessary to enable information dissemination and risk communication that consider individuals’ health literacy and beliefs. In this study, we exploratorily characterised segments based on individual health literacy and beliefs regarding COVID-19 among the Japanese during the early stage of the COVID-19 pandemic, and investigated whether infection prevention/risk-taking behaviours and fear of COVID-19 differed among these segments.

**Methods:** In this study, we conducted two web-based longitudinal surveys in Japan (PHASE 1, 1–30 November 2020, 6,000 participants; PHASE 2, 1–31 December 2020, 3,800 participants). We characterised segments of the target population using cluster analysis on health literacy and beliefs regarding COVID-19 obtained in PHASE 1. We further investigated the associations between the clusters and infection prevention/risk-taking behaviours and fear of COVID-19, obtained from PHASE 2.

**Results:** Five clusters were identified: ‘Calm/hoax denial’, ‘Hoax affinity/threat denial’, ‘Minority/indifference’, ‘Over vigilance’, and ‘Optimism’. There were significant differences in infection prevention/risk-taking behaviours and fear of COVID-19 among the five clusters. The belief in susceptibility to infection, rather than affinity for hoaxes and conspiracy theories, was coherently associated with infection prevention/risk-taking behaviours and fear of infection across clusters. This study provides foundational knowledge for creating segment-specific public messages and developing interactive risk communication to encourage infection prevention behaviours.

## Introduction

The coronavirus disease 2019 (COVID-19) pandemic, which began in 2020, has led to the promotion of behavioural regulations and infection prevention actions worldwide owing to its high infectivity and fatality rate. In the early stage of the COVID-19 pandemic, laws and penalties were often used to regulate behaviour among citizens in many countries; however, as of January 2023, there has been a shift away from the mandatory regulation of behaviours and more toward citizens’ autonomy in infection control. As COVID-19 continues to spread worldwide, it is important to identify strategies that effectively promote these voluntary prevention measures. In contrast, the Japanese government has requested citizens to adopt prevention behaviours and refrain from economic activities as from the early stage of the pandemic. These requests are voluntary rather than legal obligations of citizens, and it is up to individuals to decide what actions they take in response to the government’s call. Therefore, risk communication that encourages infection control based on autonomy, as Japan has been promoting, is becoming increasingly important worldwide.

Segment-specific risk communication about health literacy (i.e., the skill of an individual to obtain, process, and understand the health information and services needed to make appropriate health decisions (Weiss 2007; World Health Organization 1998) and beliefs is known to be effective in promoting health-related behaviour. Ishikawa et al. reported that participation rates in breast cancer screening increased by providing segment-specific information after categorising the target population into three segments based on their beliefs about cancer and its screening (Ishikawa et al. 2012).

Takemura et al. conducted a survey of optimistic or pessimistic perceptions about the probability of contracting COVID-19 and emphasized the importance of segment-specific and tailor-made risk communication amid the pandemic (Takemura et al. 2022). In order to promote infection prevention behaviours during COVID-19, it is expected to characterise segments based on individuals’ health literacy and beliefs regarding COVID-19, implementing risk communication according to these segments. Amid an infectious pandemic, risk communication aims to change behaviour by providing information. Even in one-way information provision aimed at behaviour change during a pandemic (United States Department of Health and Human Services & Prevention 2018), understanding the characteristics of the segments would be useful in constructing public messages based on the diversity of the target population. Furthermore, understanding these characteristics would be helpful in developing interactive risk communication tailored to relevant sub-groups. However, while previous studies have reported the factors associated with COVID-19 infection prevention behaviours, such as demographic factors (e.g., age, gender) (Muto et al. 2020; Pampel et al. 2010), sociodemographic factors (e.g., perception of infection risk, personality, and norms) (Bruine de Bruin & Bennett 2020; Nakayachi et al. 2020; Qian & Yahara 2020), and knowledge and information sources (Batra et al. 2021; Uchibori et al. 2022), there have been no attempts to characterise such segments based on health literacy and beliefs regarding COVID-19 or to study the relationship between segments and infection prevention behaviour.

We thus conducted two web-based longitudinal surveys with two objectives. First, we aimed to characterise segments based on health literacy and beliefs regarding COVID-19 in the first phase (PHASE 1) using an exploratory cluster analysis. We then investigated the associations between these segments obtained in PHASE 1 and infection prevention/risk-taking behaviours and the fear of COVID-19, which were assessed in the second phase (PHASE 2).

## Materials & Methods

### Ethics

This study was approved by Osaka University Graduate School of Human Sciences Research Ethics Committee (20095).

### Study Design

We conducted longitudinal questionnaire surveys on the web during two periods: 1–30 November 2020 (PHASE 1), and 1–31 December 2020 (PHASE 2).

### Participants

Participants were individuals living in Japan who had registered with Cross Marketing Inc. Cross Marketing includes over 5.4 million panellists (as of 2022) and is the largest company in its field in Japan. Participation in the survey was voluntary, and participants received ‘points’ that could be redeemed for products. This provided an incentive for participation in the survey regardless of the individuals’ interest in the survey topic.

In PHASE 1, 6,000 survey participants were recruited from monitors, aged 18–79 years. The participants were selected in terms of age (18–29, 30–39, 40–49, 50– 59, 60–69, or 70–79 years), sex (male or female), and residential area (urban or non-urban) to match their actual compositions in Japan. Target numbers were set for each of the above variables, and the survey was conducted until the target number was reached. Next, in PHASE 2, we conducted a continuous survey in which all participants from PHASE 1 were invited to participate. The target number of participants in PHASE 2 was set at 3,800 because of our budget limitation.

Inappropriate respondents in both surveys were excluded through an instructional manipulation check (Miura & Kobayashi 2019). The sex and age of the participants in PHASES 1 and 2 were as follows:

PHASE 1: Male = 3,000, female = 3,000; mean age = 49.4, standard deviation (SD) = 16.6

PHASE 2: Male = 1,969, female = 1,831; mean age = 51.7, SD = 16.0.

### Survey items

The two web surveys included the following concepts and factors. The details of the questionnaires are described in Appendix 1-a.

### PHASE 1: Health literacy and beliefs regarding COVID-19—susceptibility to infection, infection control, hoax, conspiracy theories, and optimism

Individuals’ thoughts on infectious diseases are related to the ideas and beliefs that arise from health literacy. As described above, health literacy refers to an individual’s skill in health information and services needed to make appropriate health decisions (Weiss 2007; World Health Organization 1998). In addition, it has recently been attributed not only to individual skills but also to the interaction between the individual and their surrounding environment. In other words, health literacy regarding the COVID-19 pandemic refers to individuals’ skills of obtaining health information and services and making behavioural decisions; these skills are influenced by the social context.

Furthermore, health beliefs vary from individual to individual. Since COVID-19 is a new and unknown disease, its transmission mechanism and characteristics have not been clarified. Therefore, how individuals obtain information and make decisions about infection prevention or risk-taking behaviours is likely to be mediated by their health beliefs.

The health belief model, one of the leading health behaviour theories, can provide important insight into people’s prevention/risk-taking behaviour during the COVID-19 pandemic. It states that the drivers of health behaviour include one’s perception of threat and the balance of advantages and disadvantages (Rosenstock 1974); this perception of threat consists of susceptibility (i.e., a feeling that there is a high probability of being infected with COVID-19) and severity (i.e., how serious the consequences would be if the individuals were infected with COVID-19). The balance of advantages and disadvantages is then tempered by the disadvantages (costs and barriers) of performing the behaviour. These are heavily influenced by individuals’ thoughts and beliefs.

We therefore created 82 items (six-point Likert scale; ranging from ‘strongly disagree’ to ‘strongly agree’) regarding the beliefs about COVID-19 based on previous studies on health literacy (Swami & Barron 2020; Taylor 2019) and mass media reports (i.e., newspapers, internet news). These included 35 items on susceptibility to infection; 21 items on infection control for COVID-19; and 26 items on hoax, conspiracy theories, and optimism about COVID-19. To ensure content validity, an expert in this field (KH) developed these items based on their own concepts and other authors confirmed the same. Furthermore, we used two items regarding belief in just deserts (i.e., a belief that the infected individual deserves to be infected (Murakami et al. 2022); six-point Likert scale ranging from ‘strongly disagree’ to ‘strongly agree’).

### PHASE 2: Infection prevention/risk-taking behaviour regarding COVID-19 and the fear of infection

The field of risk research has played an important role in disasters, infectious diseases, and other calamities that require individual-level to national-level measures.

Individuals’ risk perception can be categorised along two axes: dread and unknown factors. In particular, the more intuitively individuals feel dread, the stronger their demand for measures (Slovic et al., 1986). Furthermore, there are biases in this risk perception, such as present bias (O’Donoghue & Rabin 1999) and normalcy bias (Omer & Alon 1994). Regarding infection prevention behaviours for COVID-19, a bias is considered to exist wherein people put off these behaviours in favour of other behaviours even though they think infection prevention is important (i.e. present bias), alongside a bias that they will not be infected (i.e. normality bias).

Therefore, ideas about infection prevention/risk-taking behaviours regarding COVID-19 were assessed among the participants in this study based on the concepts of present bias and normality bias. Furthermore, fear of COVID-19 was included in the survey items. We originally created 18 items related to infection prevention/risk-taking behaviours regarding COVID-19 (seven-point Likert scale ranging from ‘strongly disagree’ to ‘strongly agree’). To ensure content validity, an expert in this field (KH) developed these items, and other authors confirmed them. To assess the fear of infection, instead of dread or unknown factors (Slovic 1986), we used the Perceived Vulnerability to Disease scale (PVD; seven-point Likert scale ranging from ‘strongly disagree’ to ‘strongly agree’) (Duncan et al. 2009; Fukukawa et al. 2014) that consisted of two subscales (i.e., perceived infectability and germ aversion) (see appendix 1-b).

### Statistical Analysis

We examined the distribution and homoscedasticity of variables and adopted a parametric test. To characterise the target segment, we first conducted three-factor analyses with the maximum likelihood method for questionnaires regarding health literacy and beliefs: susceptibility to infection for COVID-19; infection control for COVID-19; and hoax, conspiracy theories, and optimism about COVID-19. Promax rotation was used for this study because we assumed correlations among the extracted factors. The number of factors was comprehensively determined based on parallel analysis (Hori 2001), the scree test, and their interpretability. Factors with high loadings (≥0.3 or ≤-0.3) were considered for factor interpretation. Factor scores were obtained from the factor loadings, which were used as feature values in the subsequent cluster analysis. The adequacy of the factors obtained was confirmed using the Kaiser-Meyer-Olkin (KMO) measure of sampling adequacy. We then used cluster analysis by the k-means method with 100 iterations to characterise the segments among participants. We examined the number of clusters from three to eight with interpretability and effect size, using one-way analysis of variance (ANOVA) with Welch’s test. Effect sizes of 0.06 and 0.14 were considered medium and large, respectively (Cohen 1988). The factor scores of the factors extracted in the above factor analyses and belief in just deserts with z-standardisation were used as variables (total 14 variables; belief in just deserts was used in the previous study (Murakami et al. 2022); Cronbach’s α in this study was 0.79). Hereafter, the clusters identified by cluster analysis are referred to as segments. Differences in health literacy and beliefs among clusters were confirmed by a one-way ANOVA with Welch’s test, and the Games-Howell test as a post-hoc test to ensure that the effect sizes were sufficiently large. The criterion for effect size was as follows: η^2^= 0.01, small; 0.06, medium; and 0.14, large (Cohen 1988). The *p* value was corrected by Bonferroni correction, that is, the *p* value was multiplied by 16, the number of factors.

Next, we conducted factor analyses using the maximum likelihood method for infection prevention/risk-taking behaviours regarding COVID-19. Factor analyses were performed separately on the questionnaire items based on the concepts of present bias and normality bias. Promax rotation was applied because we assumed that there were correlations among the factors extracted. As in the above factor analyses, the number of factors was determined based on parallel analysis (Hori 2001), the scree test, and their interpretability. Cronbach’s α was also calculated to confirm reliability. We calculated the mean values of the items related to the factors obtained as well as the mean values related to the fear of infection (two variables: perceived infectability and germ aversion, in accordance with the previous study (Duncan et al. 2009; Fukukawa et al. 2014); Cronbach’s α in this study was 0.77 for perceived infectability and 0.76 for germ aversion) (a total of five factors). Finally, we conducted a one-way ANOVA with Welch’s test, and Games-Howell test as a post-hoc test, to examine the differences in these factors among clusters. The *p* value was corrected by Bonferroni correction, that is, the *p* value was multiplied by five, the number of factors.

We used SPSS (IBM SPSS, Chicago, Illinois, U.S.) version 28.0 for all analyses except the parallel analysis (Hori 2001). All *p*-values less than 0.05 were considered significant.

## Results

### Factor analyses for health literacy and beliefs and characterisation of segments using cluster analysis (PHASE 1)

#### Factor analyses for health literacy and beliefs

Regarding health literacy and beliefs about one’s susceptibility to infection with COVID-19 (35 items), five factors were obtained through factor analysis (Table 1). An adequate value in the KMO measure of sampling adequacy (=0.95) was shown. The first factor was characterised by items such as ‘People with pre-existing (underlying) diseases are more likely to be severely ill’ and ‘Elderly people are more prone to severe illness’, which we named *‘General ease of infection’*. The second factor was named *‘Extreme likelihood of infection’* because of the high factor loadings of items such as ‘Infectious by airborne transmission’, ‘Infectious by train’, and ‘transmitted from animals to humans’. Similarly, the third, fourth, and fifth factors were characterised by items such as ‘The current probability of death from infection with the new coronavirus in Japan is very low, about 1/10 million’, ‘Infections occur during nightlife (bars, clubs, host clubs, etc.)’, and ‘Young people in their 20s and 30s are spreading the novel coronavirus’, respectively; therefore, we named them *‘Low perception of infection threat’, ‘Ease of infection at dinners and parties’*, and *‘Ease of infection among young people’*, respectively.

**Table 1:**
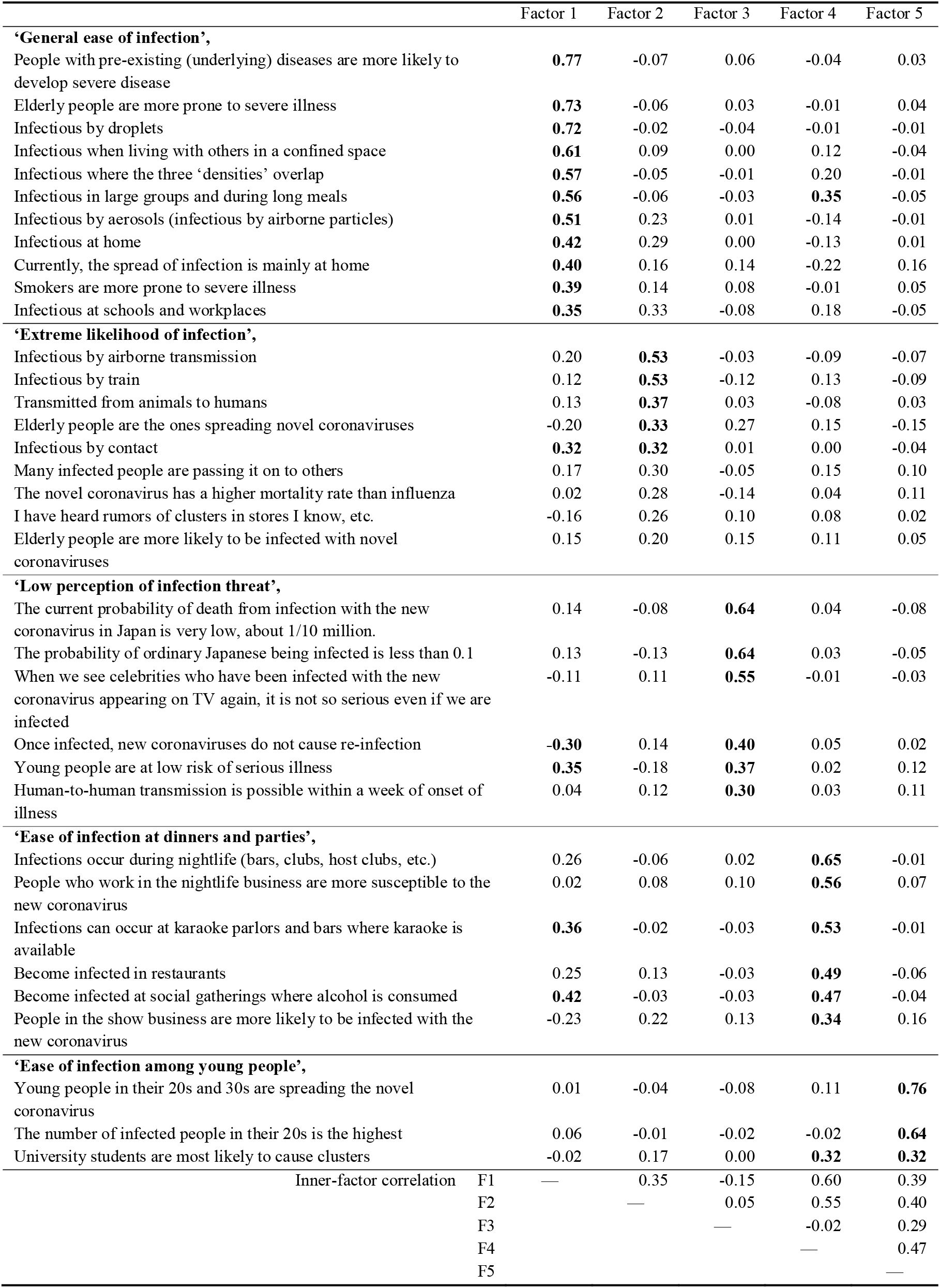
Factor loadings for health literacy and beliefs related to susceptibility to COVID-19 infection. *Factor loadings with an absolute value of 0.3 or higher are shown in bold. Items with the highest factor loadings were sorted. *The Kaiser-Meyer-Olkin measure of sampling adequacy = 0.95

Regarding health literacy and beliefs related to infection control (21 items), four factors were extracted (Table 2). There was an adequate value of the KMO measure of sampling adequacy (0.83). The first factor was characterized by items such as ‘If the polymerase chain reaction (PCR) test is negative, there is no need to worry about new coronavirus disease at all’, ‘If you take an antibody test, you do not need to take a PCR test’, and ‘Infection can be completely prevented with measures such as masks and face shields’, which we named *‘Excessive efficacy of infection control measures’*. Since the second factor showed high factor loadings of items such as ‘Routine ventilation is necessary’ and ‘vaccine can prevent severe illness after infection’, we named it *‘Efficacy of vaccines and infection control’*. Similarly, since the third and fourth factors were characterised as ‘PCR testing is intentionally suppressed’ and ‘Avigan is ineffective’, respectively, we named them *‘Dissatisfaction of PCR testing system and vaccines’* and *‘Inefficacy of therapeutic drugs’, respectively*.

**Table 2:** Factor loadings for health literacy and beliefs related to infection control for COVID-19. * Factor loadings with an absolute value of 0.3 or higher are shown in bold. Items with the highest factor loadings were sorted. *The Kaiser-Meyer-Olkin measure of sampling adequacy = 0.83

|  | Factor 1 | Factor 2 | Factor 3 | Factor 4 |
| --- | --- | --- | --- | --- |
| **‘Excessive efficacy of infection control measures’,** |  |  |  |  |
| If the PCR test is negative, there is no need to worry about the new coronavirus disease at all | **0.68** | -0.22 | 0.00 | 0.09 |
| If you take an antibody test, you do not need to take a PCR test | **0.65** | -0.19 | -0.07 | 0.04 |
| Infection can be completely prevented with measures such as masks and face shields | **0.55** | -0.02 | 0.02 | 0.02 |
| Influenza vaccine can be expected to prevent severe cardiovascular disease in new coronaviruses | **0.47** | 0.10 | 0.07 | -0.02 |
| PCR testing is the only correct test for novel coronavirus disease | **0.46** | 0.06 | 0.12 | 0.10 |
| Supine therapy (treatment with time spent lying on the stomach) is effective in the treatment of severely ill patients | **0.44** | 0.03 | 0.07 | 0.00 |
| Gargles (e.g., Isodine) are effective | **0.42** | 0.08 | 0.04 | -0.07 |
| Sodium hypochlorite is effective against novel coronaviruses | **0.35** | 0.16 | 0.06 | -0.08 |
| Treatment methods have been established to some extent in the medical field, preventing severe cases | **0.32** | 0.26 | -0.06 | -0.07 |
| Summer involves less susceptibility to novel coronavirus, while winter involves more susceptibility to novel coronavirus | 0.25 | 0.18 | 0.10 | 0.00 |
| **‘Efficacy of vaccines and infection control’,** |  |  |  |  |
| Routine ventilation is necessary | **-0.31** | **0.59** | 0.19 | 0.09 |
| Vaccine can prevent severe illness after infection | 0.11 | **0.51** | 0.01 | 0.07 |
| Vaccines for novel coronavirus are being researched and developed at a faster pace than usual | -0.08 | **0.50** | 0.04 | -0.08 |
| Japan’s PCR testing practices are correct | 0.19 | **0.50** | **-0.43** | 0.14 |
| Vaccine is effective in preventing the onset of disease after infection | 0.28 | **0.33** | 0.04 | 0.09 |
| **‘Dissatisfaction of PCR testing system and vaccines’,** |  |  |  |  |
| PCR testing is intentionally suppressed | 0.14 | -0.12 | **0.64** | -0.01 |
| PCR testing is not readily available | -0.02 | 0.13 | **0.61** | 0.10 |
| PCR testing should be performed on all | 0.06 | 0.08 | **0.50** | 0.07 |
| Vaccine for the novel coronavirus has serious side effects | 0.10 | 0.09 | **0.33** | 0.14 |
| **‘Inefficacy of therapeutic drugs’,** |  |  |  |  |
| Avigan is ineffective | 0.11 | 0.07 | 0.18 | **0.50** |
| Drugs such as Avigan, Lemdecivir, and steroids have been used in severely ill patients and are effective | 0.20 | **0.37** | 0.04 | **-0.46** |
| Inner-factor correlation |  |  |  |  |
| F 1 | — | 0.29 | 0.03 | -0.08 |
| F2 |  | — | 0.25 | -0.34 |
| F3 |  |  | — | -0.12 |
| F4 |  |  |  | — |

Regarding health literacy and beliefs related to hoaxes, conspiracy theories, and optimism for COVID-19 (26 items), four factors were extracted (Table 3). The KMO measure of sampling adequacy showed an adequate value (0.90). The first, second, third, and fourth factors showed high factor loadings for items such as ‘5G radio waves worsen coronavirus symptoms’, ‘The number of patients is increasing, nearly causing a medical collapse’, ‘It is a Chinese conspiracy’, and ‘Since July 2020, novel coronaviruses have attenuated’, respectively. We therefore named these factors *‘Hoax/conspiracy beliefs’, ‘Large social impact beliefs’, ‘China-originated beliefs’*, and *‘Optimism’*, respectively.

**Table 3:**
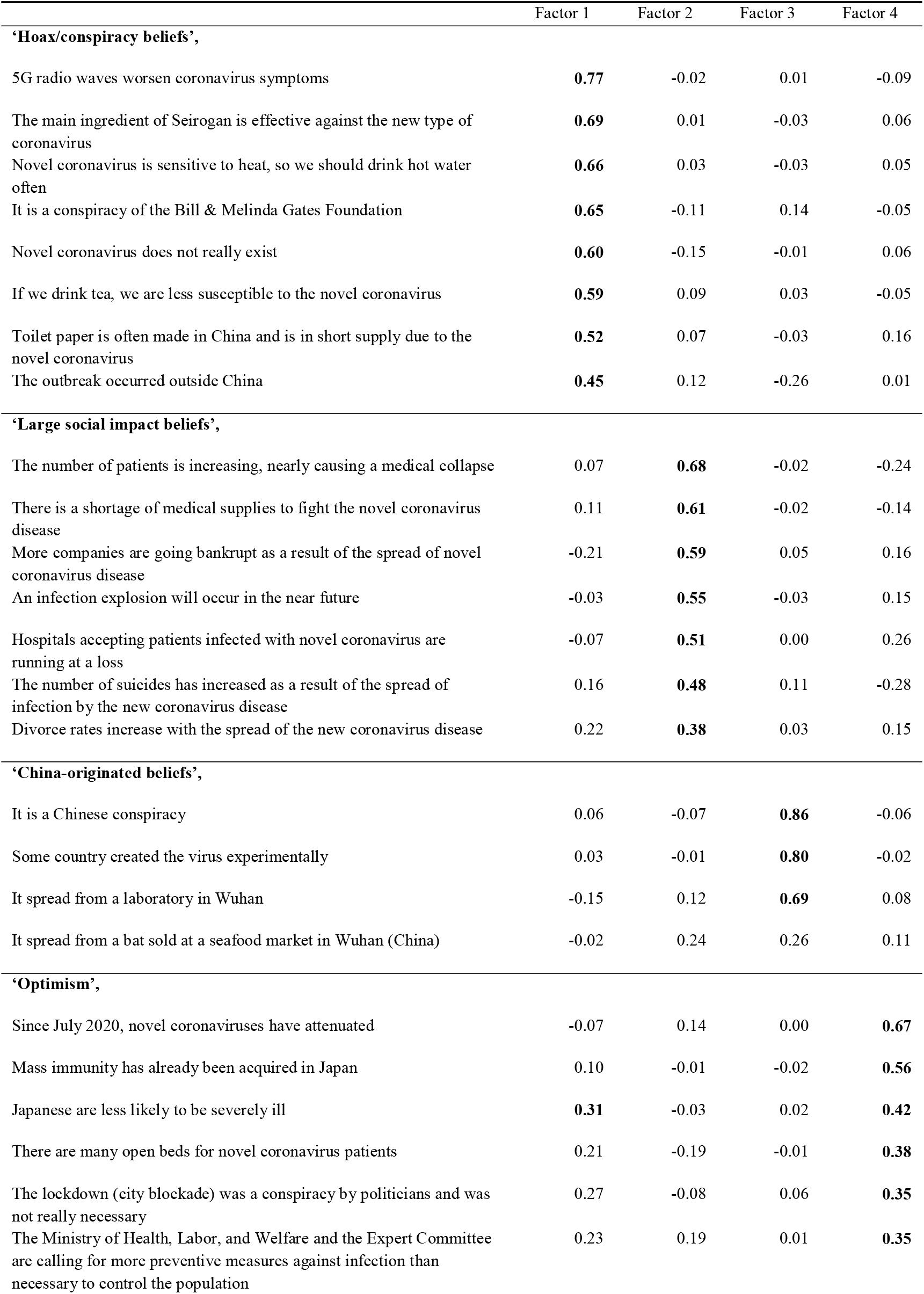
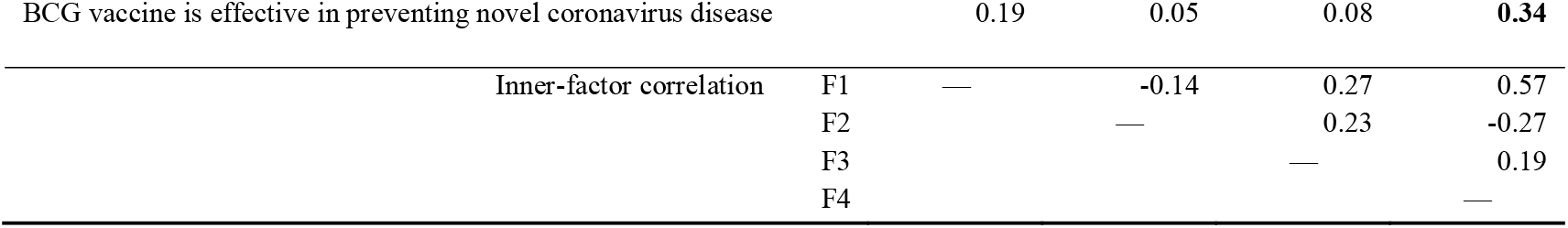
Factor loadings for health literacy and beliefs related to hoax, conspiracy theories, and optimism regarding COVID-19. * Factor loadings with an absolute value of 0.3 or higher are shown in bold. Items with the highest factor loadings were sorted. *The Kaiser-Meyer-Olkin measure of sampling adequacy = 0.90

|  | Factor 1 | Factor 2 | Factor 3 | Factor 4 |
| --- | --- | --- | --- | --- |
| **‘Hoax/conspiracy beliefs’,** |  |  |  |  |
| 5G radio waves worsen coronavirus symptoms | **0.77** | -0.02 | 0.01 | -0.09 |
| The main ingredient of Seirogan is effective against the new type of coronavirus | **0.69** | 0.01 | -0.03 | 0.06 |
| Novel coronavirus is sensitive to heat, so we should drink hot water often | **0.66** | 0.03 | -0.03 | 0.05 |
| It is a conspiracy of the Bill & Melinda Gates Foundation | **0.65** | -0.11 | 0.14 | -0.05 |
| Novel coronavirus does not really exist | **0.60** | -0.15 | -0.01 | 0.06 |
| If we drink tea, we are less susceptible to the novel coronavirus | **0.59** | 0.09 | 0.03 | -0.05 |
| Toilet paper is often made in China and is in short supply due to the novel coronavirus | **0.52** | 0.07 | -0.03 | 0.16 |
| The outbreak occurred outside China | **0.45** | 0.12 | -0.26 | 0.01 |
| **‘Large social impact beliefs’,** |  |  |  |  |
| The number of patients is increasing, nearly causing a medical collapse | 0.07 | **0.68** | -0.02 | -0.24 |
| There is a shortage of medical supplies to fight the novel coronavirus disease | 0.11 | **0.61** | -0.02 | -0.14 |
| More companies are going bankrupt as a result of the spread of novel coronavirus disease | -0.21 | **0.59** | 0.05 | 0.16 |
| An infection explosion will occur in the near future | -0.03 | **0.55** | -0.03 | 0.15 |
| Hospitals accepting patients infected with novel coronavirus are running at a loss | -0.07 | **0.51** | 0.00 | 0.26 |
| The number of suicides has increased as a result of the spread of infection by the new coronavirus disease | 0.16 | **0.48** | 0.11 | -0.28 |
| Divorce rates increase with the spread of the new coronavirus disease | 0.22 | **0.38** | 0.03 | 0.15 |
| **‘China-originated beliefs’,** |  |  |  |  |
| It is a Chinese conspiracy | 0.06 | -0.07 | **0.86** | -0.06 |
| Some country created the virus experimentally | 0.03 | -0.01 | **0.80** | -0.02 |
| It spread from a laboratory in Wuhan | -0.15 | 0.12 | **0.69** | 0.08 |
| It spread from a bat sold at a seafood market in Wuhan (China) | -0.02 | 0.24 | 0.26 | 0.11 |
| **‘Optimism’,** |  |  |  |  |
| Since July 2020, novel coronaviruses have attenuated | -0.07 | 0.14 | 0.00 | **0.67** |
| Mass immunity has already been acquired in Japan | 0.10 | -0.01 | -0.02 | **0.56** |
| Japanese are less likely to be severely ill | **0.31** | -0.03 | 0.02 | **0.42** |
| There are many open beds for novel coronavirus patients | 0.21 | -0.19 | -0.01 | **0.38** |
| The lockdown (city blockade) was a conspiracy by politicians and was not really necessary | 0.27 | -0.08 | 0.06 | **0.35** |
| The Ministry of Health, Labor, and Welfare and the Expert Committee are calling for more preventive measures against infection than necessary to control the population | 0.23 | 0.19 | 0.01 | **0.35** |

|  |  |  |  |  |  |
| --- | --- | --- | --- | --- | --- |
| BCG vaccine is effective in preventing novel coronavirus disease |  | 0.19 | 0.05 | 0.08 | **0.34** |
| Inner-factor correlation | F1 | — | -0.14 | 0.27 | 0.57 |
|  | F2 |  | — | 0.23 | -0.27 |
|  | F3 |  |  | — | 0.19 |
|  | F4 |  |  |  | — |

#### Characterisation of segments using cluster analysis on health literacy and beliefs

We conducted cluster analysis with the k-means method to characterise the participants as per health literacy and beliefs. We examined the number of clusters from three to eight with its interpretability. Finally, five clusters were adopted in this study. Table 4 shows the differences in health literacy and beliefs regarding COVID-19 among the five clusters. There were significant differences among the five clusters for all factors (*p* < 0.001). The effect sizes of η^2^ were judged as large (≥ 0.14) for all items. Furthermore, there were significant differences in the factors among clusters according to the results of the post-hoc test.

**Table 4:**
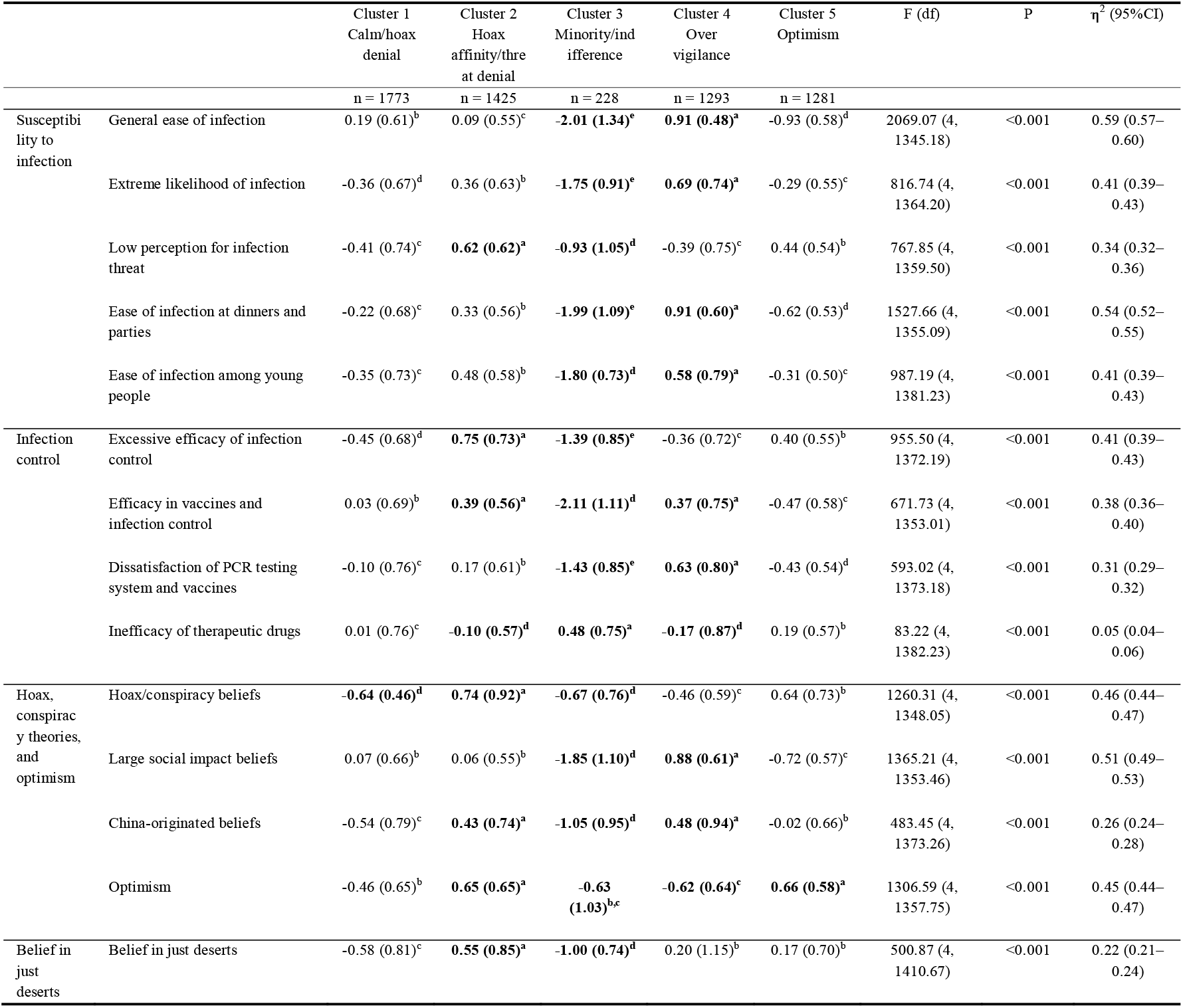
Differences in health literacy and beliefs about COVID-19 among the five clusters. *Different letters represent significant differences (P < 0.05). Higher numbers are in the alphabetical order. The highest and lowest groups are highlighted in bold font.

<u>Cluster 1</u> showed intermediate values for almost all factors among the five clusters, but it showed the lowest value only for ‘Hoax/conspiracy beliefs’. We therefore named this cluster **‘Calm/hoax denial’** (n = 1,773). <u>Cluster 2</u> was the cluster with the highest group values for ‘*Low perception of infection threat’, ‘Excessive efficacy of infection control’, and ‘Efficacy of vaccines and infection control’;* lowest group values for *‘Inefficacy of therapeutic drugs’;* and highest or second highest values for almost all factors of hoax, conspiracy beliefs, and optimism among the five clusters. Furthermore, this cluster showed the highest value for ‘Belief in just deserts’ among the five clusters. We therefore named this cluster **‘Hoax affinity/threat denial’** (n = 1,425). <u>Cluster 3</u> showed a unique profile, that is, it showed the lowest group values for almost all factors. It only showed the highest value for *‘Inefficacy of therapeutic drugs’* among the five clusters. We therefore named this cluster **‘Minority/indifference’** (n = 228). <u>Cluster 4</u> showed the highest group values for *‘General ease of infection’, ‘Extreme likelihood of infection’, ‘Ease of infection at dinners and parties’, ‘Ease of infection among young people’, ‘Efficacy of vaccines and infection control’, ‘Dissatisfaction of PCR testing system and vaccines’, ‘Large social impact beliefs’*, and *‘China-originated beliefs’;* and the lowest values for *‘Inefficacy of therapeutic drugs’* and *‘Optimism’*. Moreover, cluster 4 showed a secondary higher value for *‘Belief in just deserts’*. Therefore, we named this cluster **‘Over vigilance’** (n = 1,293). <u>Cluster 5</u> had the highest value for *‘Optimism’*. In addition, it showed secondary higher group values for *‘Low perception of infection threat’, ‘Inefficacy of therapeutic drugs’, ‘Hoax/conspiracy beliefs’, ‘China-originated beliefs’*, and *‘Belief in just deserts’*. The cluster showed secondary lower group values for *‘General ease of infection’, ‘Ease of infection at dinners and parties’*, and *‘Dissatisfaction of PCR testing system and vaccines’*. We therefore named this cluster **‘Optimism’** (n = 1,281).

### Factor analyses for infection prevention/risk-taking behaviours regarding COVID-19 and their differences among clusters (PHASE 2)

#### Factor analyses for infection prevention/risk-taking behaviours regarding COVID-19

Through factor analysis using questionnaire items based on present bias, one factor was extracted. This factor was named ‘*lack of infection prevention behaviour*’ (α = 0.82), with a consideration of the following items: ‘I am aware of the risk of infection, but I may go to a drinking party if invited’, ‘On occasions when eating or drinking with friends, if I take off my mask, I often don’t put it back on until I leave’, ‘I sometimes go to work or school even though I don’t feel as well as usual’, ‘Sometimes, I have to take off my mask at karaoke because it’s hard to sing’, ‘I sometimes eat without washing my hands and gargling’, ‘I am aware of the risk of infection, but the tourist attractions are less crowded than usual, so I tend to go on trips’, ‘Sometimes, I accidentally talk with my mask off while eating’, and ‘When I get together with friends, I tend to stay in restaurants for a long time’.

Regarding the questionnaire items based on normalcy bias, two factors were extracted (Table 5). The first factor was characterised by items such as ‘I think, “ It’s probably safe to go out for a drink today” ‘. We therefore named it *‘Acceptance of infection risk behavior’* (α = 0.83). The second factor consisted of four items, such as ‘Compared to others around me, I think I am more likely to be safe because I take better precautions against infection’ and ‘No one close to me has tested positive, so I am naturally not infected with coronavirus’. We named it *‘Sense of uninfected efficacy*’ (α = 0.72). We found sufficient consistency for all three factors.

**Table 5:**
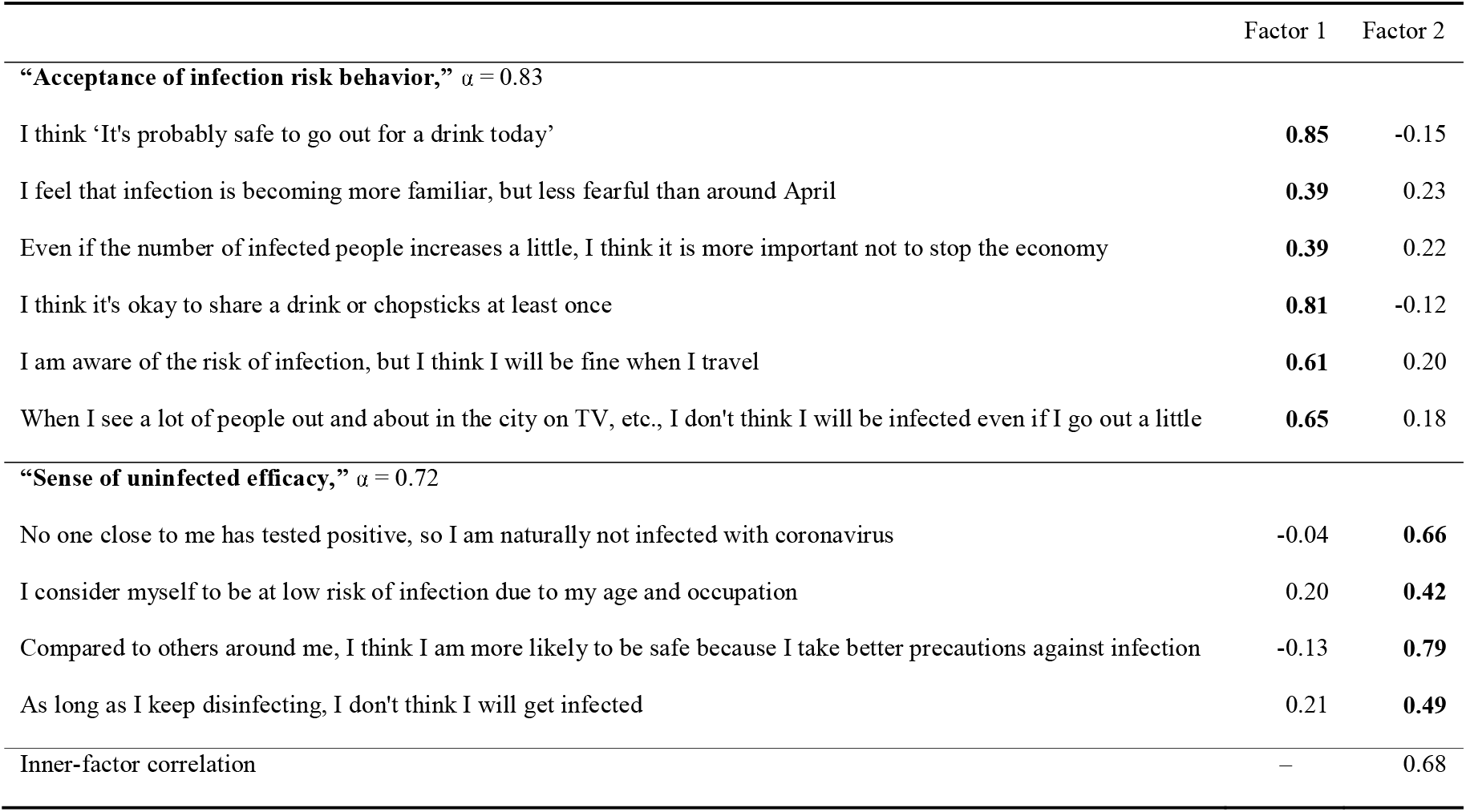
Factor pattern matrix for infection prevention/risk-taking behaviours regarding COVID-19 (normalcy bias). *Exhibited with factor loadings of 0.3 or higher in bold and sorted by the factor with the highest factor loading.

#### Differences in infection prevention/risk-taking behaviours regarding COVID-19 and the fear of infection among clusters

We found significant differences in all factors for infection prevention/risk-taking behaviours regarding COVID-19 and the fear of infection, among the five clusters: *‘Lack of infection prevention behaviour’, ‘Acceptance of infection risk behaviour’, ‘Sense of uninfected efficacy’, ‘Perceived infectability’*, and *‘Germ aversion’* (*p* < 0.001 for all factors; Table 6). In particular, *‘Acceptance of infection risk behaviour’, ‘Sense of uninfected efficacy’*, and *‘Germ aversion’* showed medium levels of effect sizes (η^2^ = 0.11, 0.07, and 0.10, respectively; Table 6).

**Table 6:** Differences in infection prevention/risk-taking behaviours regarding COVID-19 and fear of infection among the five clusters. *Different letters represent significant differences (P < 0.05). Higher numbers are in the alphabetical order. The highest and lowest groups are highlighted in bold font.

| | Cluster 1<br>Calm/hoax<br>denial | Cluster 2 Hoax<br>affinity/ threat<br>denial | Cluster 3<br>Minority/<br>indifference | Cluster 4 Over<br>vigilance | Cluster 5<br>Optimism | F (df) | P | $\eta^2$ (95%CI) |
| --- | --- | --- | --- | --- | --- | --- | --- | --- |
|  | n = 1197 | n = 907 | n = 118 | n = 856 | n = 722 |  |  |  |
| Lack of infection<br>prevention behavior | 2.18 (1.01)<sup>c</sup> | 2.32 (0.98)<sup>b</sup> | **2.34 (1.42)<sup>a,b,c</sup>** | **1.94 (0.92)<sup>d</sup>** | **2.56 (1.10)<sup>a</sup>** | 40.09 (4, 732.81) | < 0.001 | 0.04 (0.03-0.05) |
| Acceptance of infection<br>risk behavior | 2.28 (0.97)<sup>c</sup> | 2.53 (0.96)<sup>b</sup> | **2.63 (1.37)<sup>a,b,c</sup>** | **1.87 (0.86)<sup>d</sup>** | **2.90 (1.09)<sup>a</sup>** | 122.81 (4, 732.44) | < 0.001 | 0.11 (0.09-0.13) |
| Sense of uninfected<br>efficacy | 2.64 (1.07)<sup>b</sup> | **3.06 (1.04)<sup>a</sup>** | **2.51 (1.23)<sup>b,c</sup>** | **2.41 (1.12)<sup>c</sup>** | **3.14 (1.00)<sup>a</sup>** | 69.99 (4, 742.45) | < 0.001 | 0.07 (0.05-0.08) |
| Perceived infectability | 4.23 (0.93)<sup>b</sup> | 4.05 (0.84)<sup>c</sup> | **3.91 (1.03)<sup>c,d</sup>** | **4.40 (1.05)<sup>a</sup>** | **3.91 (0.81)<sup>d</sup>** | 34.00 (4, 743.81) | < 0.001 | 0.04 (0.02-0.05) |
| Germ aversion | 5.14 (0.97)<sup>b</sup> | 5.23 (0.87)<sup>b</sup> | **4.52 (1.19)<sup>c</sup>** | **5.71 (0.91)<sup>a</sup>** | **4.81 (0.89)<sup>c</sup>** | 112.83 (4, 738.61) | < 0.001 | 0.11 (0.09-0.13) |

From the results of the post-hoc test, we found significant differences in the factors among clusters. **Calm/hoax denial** (n = 1197) showed a moderate profile among the five clusters; that is, the values took the second or third place among group values for all factors regarding infection prevention/risk-taking behaviours and perceived vulnerability among the clusters. **Hoax affinity/threat denial** (n = 907) showed the highest group values for *‘Sense of uninfected efficacy’*. **Minority/indifference** (n = 118) showed the highest group value for ‘*Lack of infection prevention behaviour*’ and ‘*Acceptance of infection risk behaviour’*; and lowest group values for *‘Sense of uninfected efficacy’*, ‘*Perceived infectability*’, and *‘Germ aversion’* among clusters. **Over vigilance** (n = 856) showed the highest group values for ‘*Perceived infectability*’ and *‘Germ aversion’;* and the lowest group values for ‘*Lack of infection prevention behaviour*’, ‘*Acceptance of infection risk behaviour’*, and *‘Sense of uninfected efficacy’* among clusters. **Optimism** (n = 722) showed the highest group values for ‘*Lack of infection prevention behaviour*’, ‘*Acceptance of infection risk behaviour’*, and *‘Sense of uninfected efficacy’; and the lowest group values for* ‘*Perceived infectability*’ and *‘Germ aversion’* among clusters.

## Discussion

To develop a foundation for effective risk communication, this study characterised segments based on COVID-19 health literacy and beliefs among the Japanese in the early stage of the COVID-19 pandemic, and investigated whether infection prevention/risk-taking behaviours and fear of infection differed among the segments. We characterised the Japanese participants into five clusters based on their health literacy and beliefs regarding COVID-19, and found that the five clusters were associated with differences in infection prevention/risk-taking behaviours and fear of infection. In particular, the effect sizes of *‘Acceptance of infection risk behaviour’* and *‘Germ aversion’* were larger than those of the other clusters; these behaviours and feelings were noteworthy for their distinctive differences among clusters.

**Calm/hoax denial** had intermediate group values for the items on health literacy and belief in PHASE 1, but it had the lowest group value only for ‘Hoax/conspiracy beliefs’. In PHASE 2, this cluster also had intermediate group values for infection prevention/risk-taking behaviours and perceived fear of infection. This cluster was the most numerous of all the clusters, which may indicate that it reflected the thinking of most Japanese participants at the time this study was conducted. **Hoax affinity/threat denial** had the highest group values for ‘*Low perception for infection threat’, ‘Excessive efficacy of infection control’, and ‘Efficacy of vaccines and infection control’;* and the lowest group value for *‘Inefficacy of therapeutic drugs’*. It also had the highest or second highest group values for almost all factors of hoax, conspiracy beliefs, and optimism among the five clusters. Furthermore, this cluster had the highest value for *‘Belief in just deserts’* among the five clusters in PHASE 1. In PHASE 2, this cluster had the highest group value for *‘Sense of uninfected efficacy’*. In other words, this cluster tended to believe in the hoax and conspiracy and had a high sense of efficacy for infection control in Japan during the study period; individuals might have believed that infection was not a threat if society was taking holistic infection control measures. It might also be suggested that if these individuals were infected as a result, they believed that they would not get what they deserve. **Minority/indifference** showed a unique profile; it showed the lowest group values for almost all factors in PHASE 1. It only showed the highest group value for *‘Inefficacy of therapeutic drugs’* among the five clusters. In PHASE 2, this cluster had the highest group value for *‘Lack of infection prevention behaviour’ and ‘Acceptance of infection risk behaviour’;* and lowest group values for *‘Sense of uninfected efficacy’, ‘Perceived infectability’*, and *‘Germ aversion’* among clusters. When we interpreted the results obtained for PHASE 1 for this cluster with the questionnaire, we suspected that this cluster was not sincere in responding to the questions. In other words, there is a possibility that the cluster analysis may have selected a group that gave low scores for all items. In PHASE 2, this cluster was considered to have a high sense of efficacy in not becoming infected and a low aversion to infection, thus having a belief in infection prevention and acceptance of risk behaviours. **Over vigilance** had the highest group values for *‘General ease of infection’, ‘Extreme likelihood of infection’, ‘Ease of infection at dinners and parties’, ‘Ease of infection among young people’, ‘Efficacy of vaccines and infection control’, ‘Dissatisfaction of PCR testing system and vaccines’, ‘Large social impact beliefs’*, and *‘China-originated beliefs’;* and the lowest values for *‘Inefficacy of therapeutic drugs’ and ‘Optimism’*. The second highest group value was for *‘Belief in just deserts’* in PHASE 1. In PHASE 2, this cluster had the highest group values for ‘*Perceived infectability’* and *‘Germ aversion’*; and the lowest group values for ‘*Lack of infection prevention behaviour’*, ‘*Acceptance of infection risk behaviour’*, and ‘*Sense of uninfected efficacy*’ among clusters. In other words, members of this cluster were overly concerned about infection, with a high aversion to it, they highly estimated the ease and risk of infection, and believed that holistic infection control measures should have been taken. **Optimism** had the highest group value for *‘Optimism’* in PHASE 1. In addition, it showed secondary higher group values for *‘Low perception of infection threat’, ‘Inefficacy of therapeutic drugs’, ‘Hoax/conspiracy beliefs’, ‘China-originated beliefs’*, and *‘Belief in Just Deserts’;* including secondary lower values for *‘General ease of infection’, ‘Ease of infection at dinners and parties’*, and *‘Dissatisfaction of PCR testing system and vaccines’*. In PHASE 2, this cluster had the highest group values for ‘*Lack of infection prevention behaviour’*, ‘*Acceptance of infection risk behaviour’*, and *‘Sense of uninfected efficacy’;* and the lowest group values for ‘*Perceived infectability*’ and *‘Germ aversion’* among clusters. In other words, this cluster had a negative attitude toward infection control, was optimistic about infection, downplayed infection prevention behaviours, accepted risk behaviours, and had a low aversion to infection.

Overall, infection prevention/risk-taking behaviours and fear were associated with clusters classified based on health literacy and beliefs regarding COVID-19. Interestingly, the attitude toward strong infection prevention behaviour was found in **Over vigilance**, which was characterised by high susceptibility to infection and infection control for COVID-19. Conversely, the **Minority/indifference** cluster, which was characterised by low susceptibility to infection and infection control, did not promote infection prevention behaviours. The cluster **Optimism** also had a somewhat moderate susceptibility to infection and infection control beliefs, and had the highest levels of optimism, lack of infection prevention/risk-taking behaviours, and acceptance of infection risk behaviours. The individuals placed some emphasis on susceptibility to infection and infection control, but were characterised by optimistic beliefs. Interestingly, although ‘*hoax/conspiracy beliefs’* and ‘*China-originated beliefs’* contrasted between ‘*Calm/hoax denial’* and ‘*Hoax affinity/threat denial*’, the differences in infection prevention behaviour between these two clusters were smaller than those among the other clusters. This suggests that infection prevention/risk-taking behaviours or the fear of infection were more in harmony with beliefs about susceptibility to infection or infection control for COVID-19 than with affinity for hoaxes and conspiracy theories. The findings in this study were consistent with those of previous studies (Dryhurst et al. 2020; Harper et al. 2021; Nomura et al. 2021) reporting a strong association between infection risk perception and infection prevention behaviours in various countries.

Health insecurity, risk perception, and the resulting infection prevention behaviours in the midst of an infectious disease pandemic are greatly influenced by health literacy, which is created by information obtained from various sources, including the media and Internet (Taylor 2019). Furthermore, it has also been reported that risk perception and infection prevention behaviours regarding COVID-19 are associated with the availability of information sources (Adachi et al. 2022; Lin et al. 2020; Uchibori et al. 2022). This suggests that several information sources are likely to shape beliefs regarding susceptibility to infection or infection control, rather than through hoaxes and conspiracy theories.

This study provides foundational findings on segment characteristics regarding health literacy and beliefs toward promoting effective infection prevention behaviours. We observed a consistent association between beliefs about susceptibility to infection (or infection control) and infection prevention behaviours as well as fear across clusters, suggesting that providing public information about susceptibility or control measures against infection would be a promising strategy in the case of unilateral information dissemination from government to citizens. This presentation of risk information is known to be fundamental in the development process of risk communication (Fischhoff 1995). Furthermore, this study yielded implications for tailor-made risk communication based on segment characteristics. For example, although individuals have an affinity for hoaxes and conspiracy theories, they may have a low sense of uninfected efficacy, as seen in the clusters of **Minority/indifference** and **Over vigilance**. This illustrates the importance of choosing the content of dialogue about COVID-19 risks according to the characteristics of the segments rather than simply interacting in terms of hoaxes and conspiracy theories. Thus, effective tailor-made risk communication should be developed, with a full consideration of associations of characteristics regarding health literacy and beliefs of the target segment with their infection prevention/risk-taking behaviours or fear.

This study had some limitations. First, this study was conducted among online monitors, who are likely to be biased by the overall Japanese population. In this study, we reduced this bias by collecting participants to match the national distribution for age, sex, and residential areas. In addition, by awarding points to respondents, we provided an incentive to encourage participation, even by those who were not interested in the survey topic. Second, this study targeted the Japanese population in the early stage of the COVID-19 pandemic; therefore, caution should be exercised in applying the findings to other regions and populations at different times. It should be noted that the two-wave surveys in this study were conducted in 2020. Japan has been implementing voluntary infection prevention measures rather than mandatory behavioural regulations since the early stages of the pandemic. Therefore, this study is significant in providing foundational knowledge on risk communication to promote infection prevention behaviours in other countries that have shifted to voluntary infection prevention measures. While caution must be exercised in its application to outside populations, the study provided valuable insights into voluntary infection prevention behaviours for future research on the development of effective risk communication. Third, based on a two-wave survey, this study examined the associations between clusters based on health literacy and beliefs as well as infection prevention/risk-taking behaviours. Despite the longitudinal study design, causality was not identified. Validation based on intervention studies, such as randomised controlled trials, is needed to assess the effectiveness of segment-based risk communication in promoting effective infection prevention behaviours.

## Conclusions

In this study, we characterised five segments based on health literacy and beliefs regarding COVID-19 in Japan in the early stage of the COVID-19 pandemic, and found that these segments were associated with infection prevention/risk-taking behaviours and fear. In particular, beliefs about susceptibility to infection were found to be coherently associated with infection prevention behaviours and fear of infection across segments, implying that providing public messages about susceptibility to infection would be a promising strategy in case of unilateral information dissemination. Furthermore, the study provided foundational findings that contribute to the development of tailor-made risk communication, taking into account differences in health literacy and beliefs regarding infectious diseases of target segment characteristics.

## Data Availability

All data produced in the present work are contained in the manuscript and supplemental material.

## Acknowledgements

We would like to thank the staff of the Hirai Laboratory, Osaka University Graduate School of Human Sciences, who were involved in this study. We would also like to thank Editage (www.editage.com) for their help with English proofreading.

